# KIT-LSTM: Knowledge-guided Time-aware LSTM for Continuous Clinical Risk Prediction

**DOI:** 10.1101/2022.11.14.22282332

**Authors:** Lucas Jing Liu, Victor Ortiz-Soriano, Javier A. Neyra, Jin Chen

## Abstract

Rapid accumulation of temporal Electronic Health Record (EHR) data and recent advances in deep learning have shown high potential in precisely and timely predicting patients’ risks using AI. However, most existing risk prediction approaches ignore the complex asynchronous and irregular problems in real-world EHR data. This paper proposes a novel approach called Knowledge-guIded Time-aware LSTM (KIT-LSTM) for continuous mortality predictions using EHR. KIT-LSTM extends LSTM with two time-aware gates and a knowledge-aware gate to better model EHR and interprets results. Experiments on real-world data for patients with acute kidney injury with dialysis (AKI-D) demonstrate that KIT-LSTM performs better than the state-of-the-art methods for predicting patients’ risk trajectories and model interpretation. KIT-LSTM can better support timely decision-making for clinicians.

## I. Introduction

Clinical risk prediction using Electronic Health Record (EHR) data provides accurate and timely individualized patient outcomes, allowing early interventions for high-risk patients and better-allocating hospital resources [1], [2]. It is particularly critical to predicting risks for patients with Acute Kidney Injury requiring Dialysis (AKI-D), a severe complication associated with a very high mortality rate for critically ill patients [3], [4].

Artificial intelligence (AI), esp. deep learning (DL) models, have drawn increasing attention to patients’ outcome predictions using temporal EHR data [5], [6]. However, due to complicated data collection procedures and strict data management, EHR data are not generally AI-ready, which hinders the adaption of AI tools directly in the clinical settings [7]– [9].

Firstly, EHR data are collected daily in hospitals for efficient patient care delivery but are usually not in ideal shape for ML/DL models [5], with temporal irregularity and asynchrony being the most common problems encountered when building ML/DL applications in clinical settings [10], [11]. Irregularity refers to the uneven time gaps between measurements of a single feature. Asynchrony refers to the unaligned measurements across multiple features. Fig. 1 shows an example of EHR data with three clinical variables (SBP, HCT, and sCr) collected in the ICU. The green and blue lines in HCT show irregular time gaps between the measurements of a single clinical parameter, while the three green lines in SBP, HCT, and sCr show unaligned observations across the three clinical variables with different measurements frequency.

**Fig. 1.**
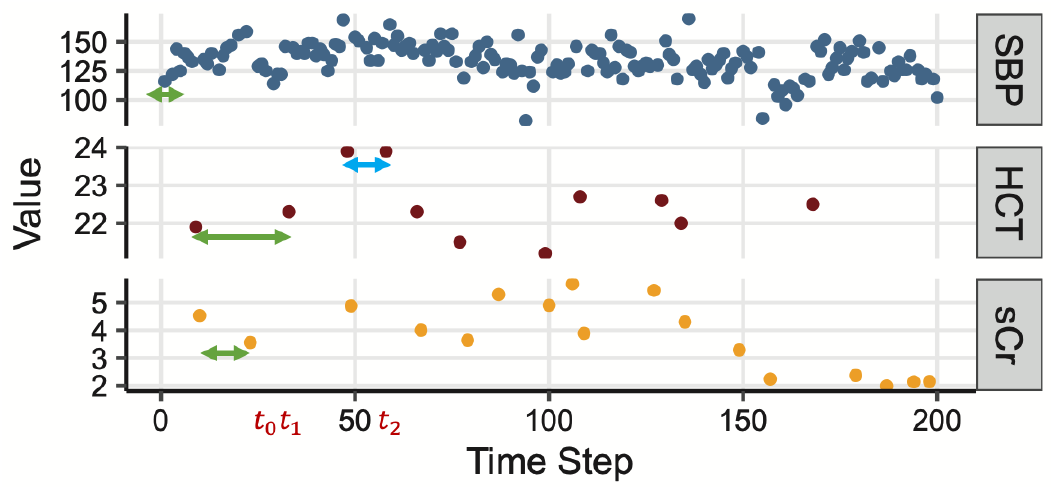
An example of real-world EHR data in the ICU. “SBP” stands for systolic blood pressure, “HCT” for Hematocrit, and “sCr” for serum creatinine. Arrows highlight irregular and asynchronous gaps between measurements.

Early DL methods ignore the irregularity and asynchrony problem. For example, the standard LSTM [12] assumes equal temporal gaps between time steps, and the original Transformer [13] uses absolute positions encoding. Recent LSTM variants have been focused on addressing the irregularity problem. T-LSTM [14] considered time elapse between patients’ visits. Phased-LSTM [15] introduced a time gate in the LSTM cell to update cell states and hidden states only if the time gate is open. Nevertheless, most of the existing LSTM models still ignore the asynchrony problem, as illustrated in Fig. 1. With recurrent neural networks (RNN), the irregularity and asynchrony problems have been addressed using missingness patterns and time elapsed between measurements. The GRU-D model updates GRU cells using missing patterns and decayed input/hidden state according to the elapsed time [16]. The BRITS model estimates missing values by introducing a complement input variable in the RNN unit when the variable is missing and also uses the hidden time decayed state of the RNN unit [17]. However, the missingness patterns are not always informative and are challenging to interpret.

Secondly, the accountability of DL models in terms of model interpretation in healthcare practice is critical for clinicians to make decisions based on the model results and rationale [18]. Model-agnostic methods, such as LIME [19] and SHAP [20], support the interpretation of any ML/DL method in a post-hoc fashion. Nevertheless, using these methods needs an extra and separate step to training the actual ML/DL models. Self-interpretable DL models such as RETAIN [21] use two-level attention scores for model interpretation, but cannot address the irregularity and asynchrony problem in EHR. ATTAIN [22] builds time-wise attention based on all/some previous cell states of LSTM plus a time-aware decay function for resolving the irregular time gaps issues. The trade-off between interpretability and prediction power invokes the development of self-interpretable ML/DL models without sacrificing prediction power, promoting a better adoption in routine uses in practical healthcare settings [23].

Another model interpretation approach uses domain-specific knowledge encoded in medical or biological ontologies databases as prior knowledge [24]. A knowledge-driven ML model that utilizes ontologies databases may gain better interpretation and potentially higher prediction power [5], [25]. Recent studies [26]–[28] have incorporated medical knowledge graphs into medical applications using translation-based graph embeddings methods [29], [30]. Moreover, medical knowledge-graph-based attention models such as GRAM [25], DG-RNN [31], and KGDAL [32] have demonstrated comparable performance as well as the power of result interpretation. Nevertheless, these methods do not embed knowledge for numerical features and lack a mechanism for handling irregular and asynchronous EHR data.

In this article, we present a Knowledge guIded Time-aware LSTM model (KIT-LSTM), which handles irregular and asynchronous time series EHR data, and uses medical ontology to guide the attention between multiple numerical clinical variables, and provides knowledge-based model interpretation.

KIT-LSTM extends LSTM with two time-aware gates and a knowledge-aware gate. The time-aware gates adjust the memory content according to two types of elapsed time, i.e., the elapsed time since the last visit for all variable streams and the elapsed time since the last measured values for each variable stream. The knowledge-aware gate uses medical ontology to guide attention between multiple numerical variables at each time step. To the best of our knowledge, KIT-LSTM is the first LSTM variant that incorporates medical ontology with the addition of two time-aware gates to guide the attention mechanism inside the LSTM cell. As a result, the proposed model provides better guidance for attention and interpretation and handles both irregular and asynchronous problems simultaneously. Our contributions are summarized as follows:

1. KIT-LSTM adds to the original LSTM cell two unique time-aware gates. The time-ware gates adjust different proportions of the LSTM cell memory contents, which address irregularity and asynchrony.
2. KIT-LSTM adds to the original LSTM cell a knowledge-aware gate. It uses the relationship between concepts learned from medical ontology to guide attention between multiple variables at each time step, and the loss function enforces the learned attention aligned with the medical ontology, enabling knowledge-based model interpretation.
3. Using EHR data, KIT-LSTM continuously and accurately predicts mortality risks in the next 24 hours for critically ill AKI-D patients in the ICU.
4. KIT-LSTM shows high robustness in subpopulation distribution shift.

## II. Method

### A. Notations

#### EHR features

A patient’s EHR data at time step *t* can be represented as a vector of clinical parameters (e.g., heart rate) denoted as 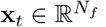, where *N*_*f*_ is the number of features.

#### Time-related features

Δ_*t*_ ∈ ℝ denotes the time elapsed since the last time step, and 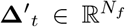 denotes the time elapsed since the last measured value of the same feature. The value of 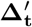 for each feature can be different since features are possibly measured at different frequencies.

#### Knowledge-related features

KIT-LSTM uses Human Phenotype Ontology (HPO) [33] as the prior knowledge to guide the model learning process. We extract ontology concepts related to the selected clinical features and call them feature concepts (e.g., “elevated systolic blood pressure” (HP:0004421) is a concept related to systolic blood pressure in HPO). In addition, we extract a concept related to the study population, i.e., “acute kidney injury” (HP:0001919), which is called the target concept. The total number of ontology concepts is *N*_*o*_ +1, where *N*_*o*_ is the number of feature concepts. Note that *N*_*o*_ is not the same as the number of features *N*_*f*_ because some features can be mapped to more than one related ontology concept, and some can only be mapped to one. All concepts are encoded as one-hot vectors as the initial embeddings denoted as 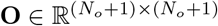.

Based on the feature values and the ontology concepts at time step *t*, we extract the physiological status denoted as 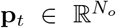. The value of *p*_*t*_ for each concept is either 0 or 1. For example, *p*_*t*_ = 1 for concept “high systolic blood pressure” means patient systolic blood pressure is greater than 130. Thresholds of all features are defined based on clinical practice and are validated by clinicians.

### B. KIT-LSTM Cell

The architecture of KIT-cell is illustrated in Fig. 2. The input to a KIT-LSTM cell consists of five components: clinical feature **x**_*t*_, elapsed time Δ_*t*_ since last time step, elapsed time 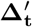 since the last measured values, physiological status **p**_*t*_, and initial concept embedding **O**. KIT-LSTM kept the original three gates (forget, input, output) from LSTM and added three additional gates (two time-aware gates and one knowledge-aware gate).

**Fig. 2.**
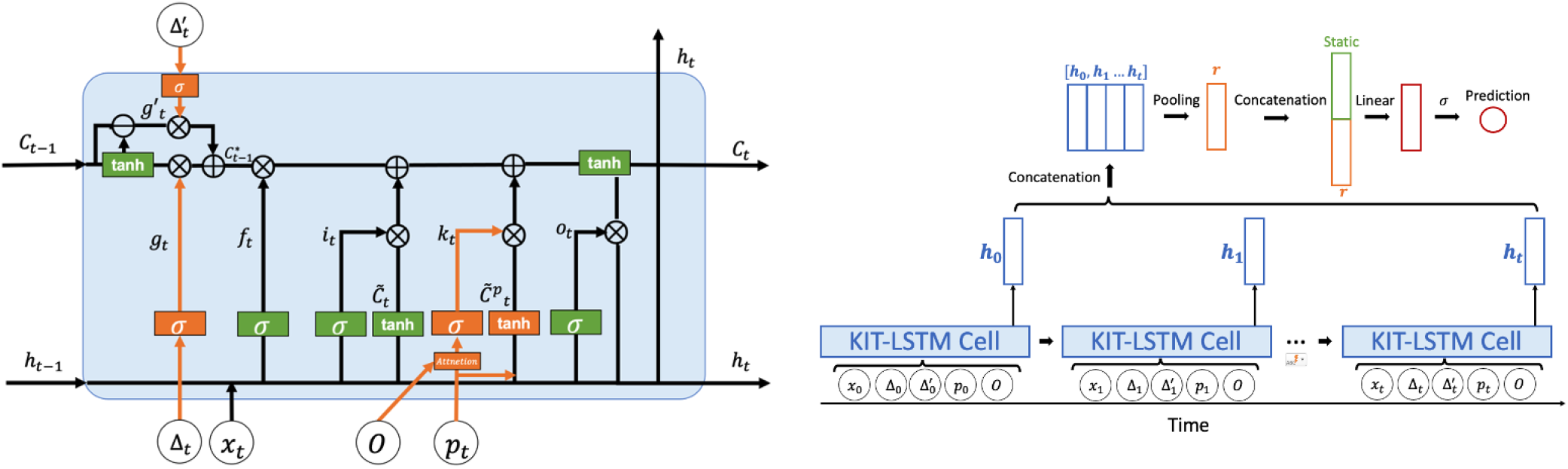
The architecture of KIT-LSTM cell (left), orange represents the unique gates in KIT-LSTM, and green represents the original gates in LSTM. The prediction layers (right) combine all the hidden states learned from KIT-LSTM and the static features, such as patient demographics, for the final prediction.

The first time gate, the long-term time-aware gate, is a time decay function that adjusts long-term memory by using the elapsed time since the last measured value of the same clinical parameter. For example, the green and blue arrows in Fig. 1 for the “HCT” indicate that the previous memory will be more likely to be discounted when the green arrow ends (*t*_1_) than the time when the blue arrow ends (*t*_2_).

The second time gate, the short-term time-aware gate, is a time decay function to adjust short-term memory. It is measures the elapsed time since the last time step (e.g, *t*_1_ *− t*_0_ in Fig. 1), similar to the time decay function in T-LSTM [14].

The long and short time-aware gates control how the previous short or long-term memories can be passed into the current memory. Intuitively, the longer the elapsed times, the less likely the long or short-term memory gate will open.

The knowledge-aware gate uses concepts embeddings and physiological features to control which feature should be paid more attention to at each time step. Intuitively, if the physiological status is abnormal and there is a strong relationship between a feature concept and the target concept, more attention will be paid to the corresponding feature.

#### Gate update

We denoted the forget gate as **f**_*t*_, the input gate as **i**_*t*_, the output gate as **o**_*t*_, the time-aware gates as **g**_*t*_ and **g**^*′*^*t*, and the knowledge-aware gate as **k**_*t*_, where **f**_*t*_, **i**_*t*_, **o**_*t*_, **g***t*, **g**^*′*^_*t*_, **k**_*t*_ *∈* ℝ^*m*^, *m* is the dimension of the hidden vectors, and *t* is the time step. The short-term time-aware gate **g**_*t*_ is updated by the elapsed time since the last time step, and the long-term time-aware gate **g**^*′*^*t* is updated by the elapsed time since last measured value for each feature. The knowledge-aware gate **k**_*t*_ is updated by the attention scores 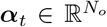 learned from the concept embeddings as well as physiological status **p**_*t*_. Gates at time step *t* are: ^1^

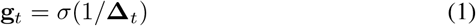

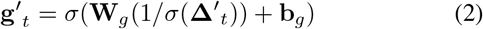

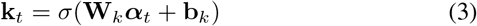

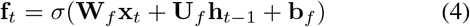

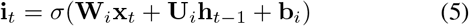

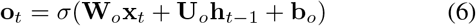

Where 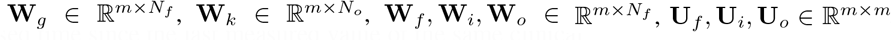, and **b**_*g*_, **b**_*k*_, **b**_*f*_, **b**_*i*_, **b**_*o*_ ∈ ℝ ^*m*^ are the learnable parameters. *σ* is a sigmoid function.

The attention score ***α***_*t*_ for each physiological feature at time *t* is computed using:

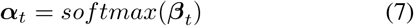

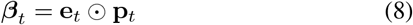

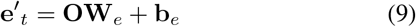

Where 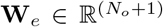 and 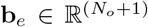 are the learnable parameters; and 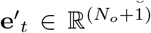 is the learned concepts embedding from initial embeddings **O** using a linear function including the feature concepts embeddings 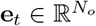 and the target concept embedding denoted as *e*_*target*_ *∈* ℝ. Note that *e*_*target*_ is not shown in above equation, but will be used in loss regularization described in Section II-C.

#### Memory cell update

Following the definition in T-LSTM [14], we extracted the short and long-term memory from the previous memory cell **C**_**t***−***1**_ *∈* ℝ^*m*^, denoted as 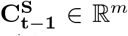 and 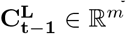 respectively. Then, the short- and long-term memory are adjusted separately by their corresponding time-aware gates **g**_*t*_ and **g**^*′*^_*t*_. In particular, the short-term memory is discounted by the elapsed time since last time step, and the long-term memory is discounted by the elapsed time since last measured values. We denote the discounted short-term and long-term memory cell as 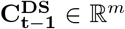 and 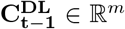 respectively. Finally, the total adjusted previous memory cell denoted as 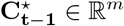 is the sum of all the discounted short-and long-term memories. Memory cells are computed as:

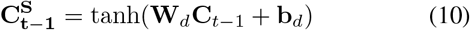

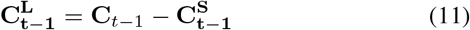

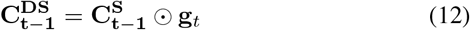

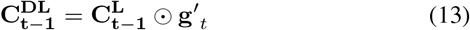

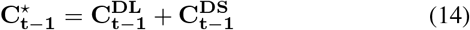

where **W**_*d*_ *∈* ℝ^*m×m*^, **b**_*d*_ *∈* ℝ^*m*^ are the learnable parameters, and *⊙* is the Hadamard product.

#### Candidates memory cells

While the original feature candidate cell, denoted as 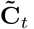, computed from the feature value **x**_*t*_ and the previous hidden state **h**_*t*_, a new candidate cell is added, denoted as 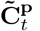, which also considers the physiological status **p**_*t*_. Candidate memory cells can be computed using:

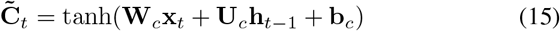

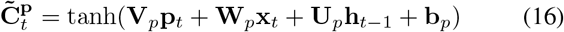

where 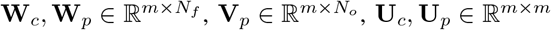 and **b**_*c*_, **b**_*p*_, *∈* ℝ^*m*^ are the learnable parameters.

#### Current memory cell and hidden state

**C**_*t*_ *∈* ℝ^*m*^ and **h**_*t*_ *∈* ℝ^*m*^ represent the current memory cell and its corresponding hidden state. **C**_*t*_ is a combination of adjusted previous memory 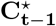 multiplied by the forget gate, the feature candidate memory multiplied by the input gate, and the physiological candidates memory multiplied by the knowledge-aware gate. The current cell and hidden state are computed using:

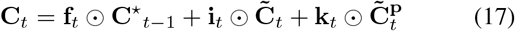

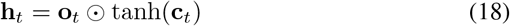

### C. Patient Outcome Prediction

#### Prediction layer

All the hidden states **h**_*t*_ are concatenated and passed into a pooling layer for taking the sum/max along time steps. The resulting hidden representation is denoted as **r** *∈* ℝ^*m*^. Then the static features (e.g, demographics) are concatenated with **r** followed by a fully connected layer with a sigmoid function for the binary prediction. The process of final prediction is shown in Fig. 2.

#### Loss function

Let the ground truth label be *y* and the predicted label be ŷ, we use the binary cross entropy as the part of the final prediction loss denoted as *L*_*pred*_. Inspired by a knowledge graph guided model KGDAL [32], where a regularization term is employed to consolidate the concept relations from medical ontology into attentions, we add a similar regularization term to ensure the relationship between the learned feature concept embeddings **e** and the target concept embedding **e**_*target*_ aligns to the observed relations in a medical ontology. Thus, the regularization term counts the discrepancy at the knowledge level, i.e. the difference between the learned concept embedding distance and the corresponding concept distance in a medical ontology. The regularization term *L*_*reg*_, cross entropy loss *L*_*pred*_, and final prediction loss *L* are:

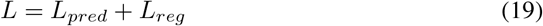

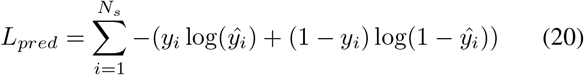

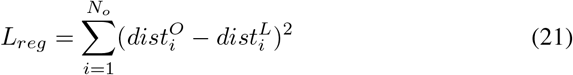

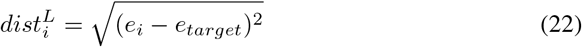

where *N*_*s*_ is the total number of samples. 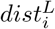 represents the distance between feature concept embedding *i* and the target embedding; similarly 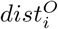 represents the distance between feature concepts *i* and the target concept in medical ontology. The observed distance can be obtained directly from the ontology graph by computing node-based distances or it can be obtained from the pre-trained initial embedding using graph embedding methods [29].

The source code of KIT-LSTM is available at https://github.com/lucasliu0928/KITLSTM.

## III. Experiment Settings

The experiment aims to continuously predict AKI-D patients’ mortality risk in their dialysis/renal replacement therapy (RRT) duration. More specifically, given any period of EHR in dialysis duration before time *T*, we will continuously predict the mortality risk at *T* + 24, i.e., 24 hours after *T*.

### A. Experiment Data

#### Patient cohort

The study population consists of 570 AKI-D adult patients admitted to ICU at the University of Kentucky Albert B. Chandler Hospital from January 2009 to October 2019. Among them, 237 (41.6%) died in hospital, and 333 (58.4%) survived. Patients were excluded if they were diagnosed with end-stage kidney disease (ESKD) before or at the time of hospital admission, were recipients of a kidney transplant, or had RRT less than 72 or greater than 2,000 hours.

#### EHR data

Data features include twelve temporal features (systolic blood pressure, diastolic blood pressure, serum creatinine, bicarbonate, hematocrit, potassium, bilirubin, sodium, temperature, white blood cells (WBC) count, heart rate, and respiratory rate) and six static features (age, race, gender, admission weight, body mass index (BMI), and Charlson comorbidity score). All outliers greater than 97.5 or lower than 2.5 percentile were excluded. Measurement frequencies vary dramatically, ranging from 0.2 to 21.7 observations per day.

#### Sample generation

To continuously predict patient’s mortality risks, we generate 30 samples from each patient’s EHR data with a random start and end time as long as the length of the sample is greater than 10 time steps where a time step refers to the time when any feature has a value. The class label of a sample is whether the patient died (positive) or survived (negative) in the next 24 hours from the end of the sample.

#### Obvious negative sample exclusion

The negative to the positive ratio in the data is 10:1 at the sample level. In such cases, ML models may be biased to the negative, resulting in so-called “good” performance. Hence, we excluded “obvious negative samples” from EHR, allowing models to focus on the more difficult cases and better balance positives and negatives in model training. This process is applied to all the compared methods to ensure fair performance comparison. The process is described as follows: 1) apply PCA [34] on the average, minimum or maximum values of all temporal and static features; 2) For each sample, compute the weighted sum of the top seven features using the squared correlation (contribution score) to the first principal component as weights; 3) determine obvious negative samples using the distribution of the weighted sum values. In total, 4,563 (or 25%) obvious negative samples were identified and excluded.

#### Training, validation and testing data

From 570 AKI-D patients, 13,333 EHR samples were extracted, including 1,456 positives and 11,877 negatives. As shown in Table I, the data were split into training (75%), validation (5%), and testing data (20%) patient-wise to ensure that the samples from the same patients only appeared in one of the three datasets.

**TABLE I.**
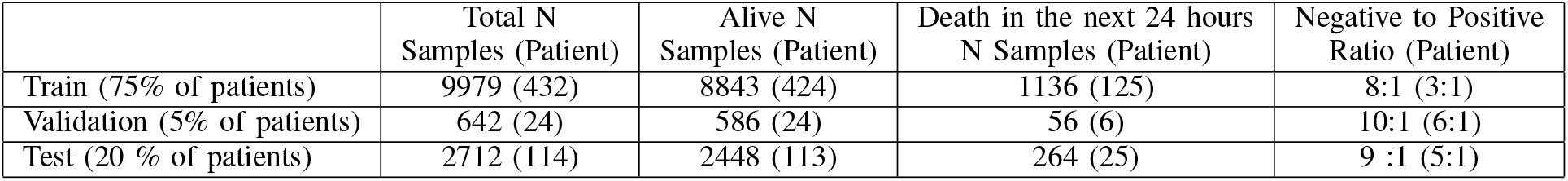
Training,Validation and Testing Data.

|  | Total N<br>Samples (Patient) | Alive N<br>Samples (Patient) | Death in the next 24 hours<br>N Samples (Patient) | Negative to Positive<br>Ratio (Patient) |
| --- | --- | --- | --- | --- |
| Train (75% of patients) | 9979 (432) | 8843 (424) | 1136 (125) | 8:1 (3:1) |
| Validation (5% of patients) | 642 (24) | 586 (24) | 56 (6) | 10:1 (6:1) |
| Test (20 % of patients) | 2712 (114) | 2448 (113) | 264 (25) | 9 :1 (5:1) |

### B. Baseline algorithms

We compared KIT-LSTM with eight existing algorithms, including two traditional ML algorithms (XGBoost and SVM) and six DL models (LSTM, T-LSTM, Phased-LSTM, RETAIN, ATTAIN, and Transformer). For all DL models, static features are concatenated with the hidden states before the prediction layer, as described in Section II-C. Moreover, all missing temporal features are imputed with the last observation carried forward (LOCF) method.

### C. Model robustness evaluation metric

Robustness is one of the most important performance metrics for clinical applications, which can be assessed using the subpopulation distribution shift approach [35], [36]. First, patient subpopulations were identified using demographics and comorbidity in EHR. Second, subpopulations at different levels of granularity were obtained using hierarchical clustering and applying thresholds at the dendrogram. Third, the clustering dendrogram and the corresponding t-SNE plot of subpopulations were visualized to identify distinct subpopulations. Fig. 3 shows four subpopulations at the second level (green) of dendrogram have distinctly different distributions.

**Fig. 3.**
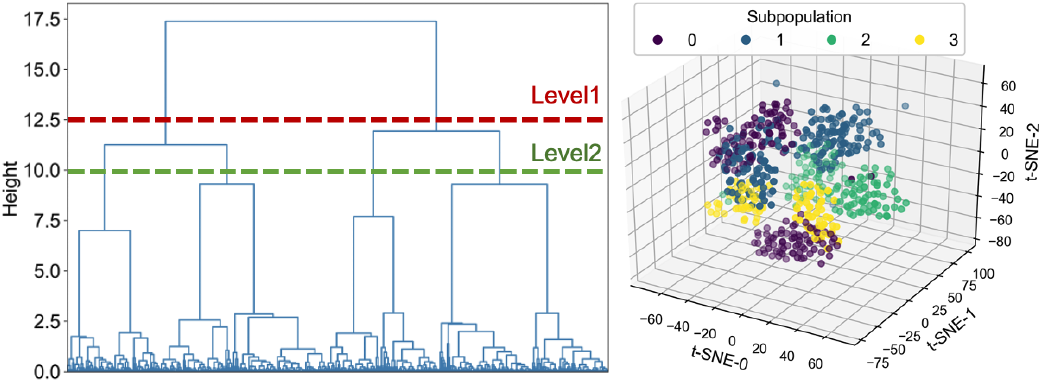
Identified subpopulation using hierarchical clustering. In the clustering dendrogram (left), the horizontal lines show two different levels of subpopulations. The resulting subpopulations at level two (right) are shown on a three-dimensional t-SNE space.

## IV. Results and Discussion

Table II shows the overall performance on both balanced and imbalanced test data (pos: neg ratio being 1:1 and 1:9) of our proposed model KIT-LSTM compared with other baseline models. KIT-LSTM has the best overall performance with the highest ROCAUC, F-3, and recall on both test data and the highest F1 on balanced data. The traditional ML method XGBoost has the highest precision on both test data and the highest accuracy on the imbalanced test data, but the scores on all other metrics are the lowest.

**TABLE II.** Overall Performance of KIT-LSTM and seven compared algorithms on balanced and imbalance test sets

|  | Balanced Test (Pos:Neg = 1:1) |  |  |  |  |  | Imbalanced Test (Pos:Neg = 1:9) |  |  |  |  |  |
| --- | --- | --- | --- | --- | --- | --- | --- | --- | --- | --- | --- | --- |
|  | ROCAUC | ACC | F-1 | F-3 | Recall | Precision | ROCAUC | ACC | F-1 | F-3 | Recall | Precision |
| XGBoost | 0.55 | 0.55 | 0.18 | 0.11 | 0.10 | **0.96** | 0.55 | **0.91** | 0.17 | 0.11 | 0.10 | **0.65** |
| SVM | 0.63 | 0.59 | 0.58 | 0.57 | 0.56 | 0.59 | 0.66 | 0.67 | 0.25 | 0.45 | 0.56 | 0.16 |
| LSTM | 0.66 | 0.64 | 0.56 | 0.48 | 0.46 | 0.72 | 0.67 | 0.80 | 0.31 | 0.42 | 0.46 | 0.24 |
| Transformer | 0.70 | 0.59 | 0.39 | 0.28 | 0.26 | 0.79 | 0.70 | 0.87 | 0.28 | 0.26 | 0.26 | 0.30 |
| T-LSTM | 0.70 | 0.63 | 0.55 | 0.47 | 0.46 | 0.70 | 0.70 | 0.77 | 0.28 | 0.41 | 0.46 | 0.20 |
| Phased-LSTM | 0.65 | **0.65** | 0.56 | 0.47 | 0.45 | 0.74 | 0.66 | 0.82 | **0.33** | 0.42 | 0.45 | 0.26 |
| RETAIN | 0.68 | 0.64 | 0.54 | 0.44 | 0.42 | 0.74 | 0.67 | 0.80 | 0.29 | 0.39 | 0.42 | 0.23 |
| ATTAIN | 0.60 | 0.55 | 0.39 | 0.30 | 0.28 | 0.61 | 0.58 | 0.76 | 0.19 | 0.26 | 0.28 | 0.14 |
| KIT-LSTM (Ours) | **0.72** | 0.64 | **0.62** | **0.58** | **0.58** | 0.67 | **0.71** | 0.71 | 0.28 | **0.47** | **0.58** | 0.18 |

To assess model robustness using subpopulation shift, we measured performance variability across subpopulations at different levels of granularity. The performance on both balanced and imbalanced test data are shown in Table III and Table IV respectively, the scores are shown in average and standard deviation (in parentheses).

**TABLE III.**
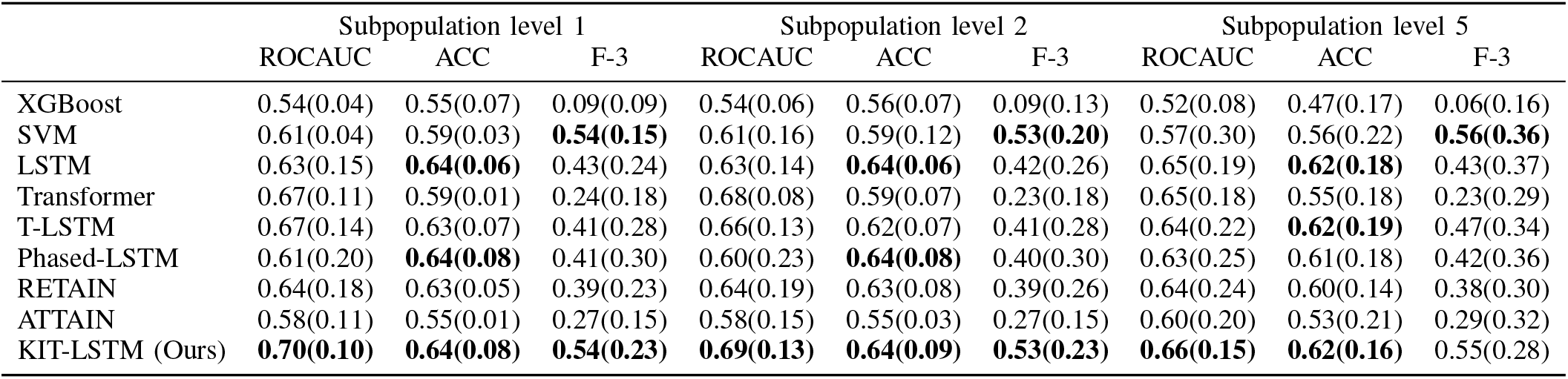
Balanced performance of KIT-LSTM and seven compared algorithms on multiple subpopulation levels (pos:neg = 1:1).

**TABLE IV.** Imbalanced performance of KIT-LSTM and compared algorithms on multiple subpopulation levels (pos:neg = 1:9).

|  | Subpopulation level 1 |  | Subpopulation level 2 |  | Subpopulation level 5 |  |
| --- | --- | --- | --- | --- | --- | --- |
|  | ROCAUC | F-3 | ROCAUC | F-3 | ROCAUC | F-3 |
| XGBoost | 0.54(0.04) | 0.09(0.08) | 0.54(0.06) | 0.09(0.13) | 0.52(0.07) | 0.06(0.16) |
| SVM | 0.64(0.04) | 0.43(0.14) | 0.63(0.15) | 0.42(0.18) | 0.63(0.26) | 0.46(0.32) |
| LSTM | 0.64(0.14) | 0.38(0.22) | 0.63(0.13) | 0.37(0.24) | **0.65(0.22)** | 0.38(0.33) |
| Transformer | 0.68(0.09) | 0.22(0.17) | **0.69(0.08)** | 0.22(0.17) | 0.63(0.22) | 0.22(0.29) |
| T-LSTM | 0.67(0.12) | 0.36(0.25) | 0.67(0.12) | 0.35(0.24) | 0.64(0.23) | 0.42(0.31) |
| Phased-LSTM | 0.62(0.20) | 0.37(0.27) | 0.61(0.22) | 0.35(0.28) | 0.63(0.28) | 0.38(0.34) |
| RETAIN | 0.63(0.18) | 0.35(0.21) | 0.63(0.19) | 0.34(0.23) | 0.63(0.26) | 0.35(0.28) |
| ATTAIN | 0.56(0.11) | 0.24(0.14) | 0.55(0.13) | 0.23(0.14) | 0.60(0.20) | 0.26(0.31) |
| KIT-LSTM (Ours) | **0.69(0.08)** | **0.44(0.20)** | **0.69(0.12)** | **0.43(0.20)** | 0.64(0.17) | **0.47(0.26)** |

Table III and Table IV show that overall KIT-LSTM out-performs all the compared methods on almost all tested subpopulations on all evaluation metrics. SVM has the same highest F-3 as KIT-LSTM on the balanced test data at level 1 and 2, but the ROCAUC of SVM is at the lower end at all levels. LSTM has the highest ROCAUC on the unbalanced data at level 5, but its F-3 on the same data is moderate.

Table III and Table IV show that the tested DL methods (except for ATTAIN) outperformed the traditional ML methods XGBoost and SVM on almost all metrics. ATTAIN has the second worst performance on balanced and unbalanced data. Phased-LSTM and RETAIN have moderate performance. Phased-LSTM has comparable accuracy on the balanced data, but its ROCAUC is on the lower end. Transformer and T-LSTM are the most competitive methods as they performed the second or third to the best on ROCAUC for all different subpopulations on both balanced and imbalanced test data. Table IV shows that Transformer has the same highest ROCAUC score as KIT-LSTM at level 2 subpopulation on the imbalanced data, but its F-3 score is at the lower end. T-LSTM maintained comparable performance on all datasets, which we considered the second-best model after KIT-LSTM.

Regarding the model robustness, we compared the variation of ROCAUC scores on subpopulations within a single level. XGBoost has the lowest standard deviation at all three subpopulation levels. Nevertheless, the average ROCAUC scores are lower than KIT-LSTM. On the other hand, KIT-LSTM has the best performance where its average ROCAUC scores are the highest, and the standard deviation of ROCAUC is the second-or the third-smallest at all three subpopulations levels.

### A. Ablation Study

We conducted an ablation study to test how each component of KIT-LSTM performed by removing several components of KIT-LSTM. The variants are:

**KIT**_*−k*_: remove knowledge-aware gate **k**_*t*_ while keeping two time-aware gates and physiological status **p**_*t*_.

**KIT**_*−kp*_: remove knowledge-aware gate **k**_*t*_ and physiological status **p**_*t*_ while keeping the two time-aware gates.

**KIT**_*−kt*_: remove knowledge-aware gate **k**_*t*_ and two time aware gates while keeping physiological status features.

Table V shows that KIT_*−kt*_ without any time-aware gates or knowledge-aware gates has the lowest performance, whereas models maintained time-aware gates (KIT_*−kp*_ and KIT_*−k*_) had better performance. The lower performance of KIT_*−kt*_ and KIT_*−k*_ indicates that the physiological status itself is not adequate for enhancing model prediction power, no matter whether it is added alone (KIT_*−kt*_) or added with time-aware gates (KIT_*−k*_).

**TABLE V.** Ablation study: Performance of KIT-LSTM variants on balanced data at level 1, 2 and 5 subpopulations.

|  | ROCAUC | Level 1<br>ACC | F-3 | ROCAUC | Level 2<br>ACC | F-3 | ROCAUC | Level 5<br>ACC | F-3 |
| --- | --- | --- | --- | --- | --- | --- | --- | --- | --- |
| $KIT_{-kp}$ | 0.65(0.15) | 0.63(0.06) | 0.44(0.26) | 0.65(0.16) | 0.63(0.06) | 0.43(0.28) | 0.62(0.24) | 0.60(0.17) | 0.40(0.37) |
| $KIT_{-kt}$ | 0.60(0.16) | 0.59(0.05) | 0.48(0.22) | 0.59(0.21) | 0.58(0.07) | 0.47(0.22) | 0.56(0.25) | 0.56(0.19) | 0.45(0.34) |
| $KIT_{-k}$ | 0.65(0.17) | 0.62(0.06) | 0.37(0.27) | 0.65(0.17) | 0.61(0.08) | 0.36(0.27) | 0.64(0.19) | 0.59(0.16) | 0.40(0.33) |
| KIT-LSTM | **0.70(0.10)** | **0.64(0.08)** | **0.54(0.23)** | **0.69(0.13)** | **0.64(0.09)** | **0.53(0.23)** | **0.66(0.15)** | **0.62(0.16)** | **0.55(0.28)** |

KIT-LSTM has the best performance for all subpopulations. Especially, the F-3 scores are 6-17% higher than the models in the ablation study, indicating that the two time-aware gates and the knowledge-aware gate have a substantial effect on improving the prediction power. In particular, when comparing KIT-LSTM with the latest baseline methods T-LSTM and ATTAIN that also used a time-aware gate, KIT-LSTM has better performance, indicating that both the long-term time-aware gate and the knowledge-aware gate play an essential role.

### B. Model Validation and Interpretation

Four survived and three dead cases were randomly selected for clinical validation. Two experienced clinicians reviewed the patient records independently while the predicted risk and true label were blinded in the validation process. For the dead cases, KIT-LSTM predicts them all with high risk, whereas the clinicians underestimate the risk for one case. For the survived cases, KIT-LSTM overestimate three case, while the clinicians overestimate one case. The results suggest that KIT-LSTM has a higher recall than precision. In clinical settings, higher recall is more important than higher precision. In this case, the high-risk patient correctly predicted by KIT-LSTM but underestimated by clinicians could cause severe clinical problems.

Attention scores ***α***_*t*_ obtained at each time step are used to interpret the behavior of KIT-LSTM. Fig. 4 illustrates the attention scores obtained from two adjacent samples of one patient. Case “SP12” with a low risk is predicted correctly by KIT-LSTM and clinician. Another case “SP2” has a relatively higher risk predicted by both KIT-LSTM and clinician. The figure shows that the attention scores for respiratory rate (RR) are high for almost all time for both SP12 and SP2, but the interpretation of attention should be different considering the opposite predicted outcome.For example, for the low-risk patient SP12, most of the RR values are steady within a mid-high normal range of 15-22 breaths per minute. Thus, the corresponding attention scores suggests that these values highly contributed to the low-risk prediction. For high-risk patient SP2, the RR values are relatively less steady than SP12. There are more lower-end values (before 10 hours) and more higher-end values (between 20 and 40 hours). Thus, the attention scores suggest these values contributed more towards high risk. For diastolic blood pressure (DBP), both attention scores and feature values show a bump along the time. However, the bump that happened earlier (far way from the prediction window) for SP12 contributes to lower risk prediction, and the bump that happened later (Closer to the prediction window) for SP2 contributions to a higher risk prediction. With both the trajectories and the attention scores, KIT-LSTM can assist clinicians in making better and timely decisions.

**Fig. 4.**
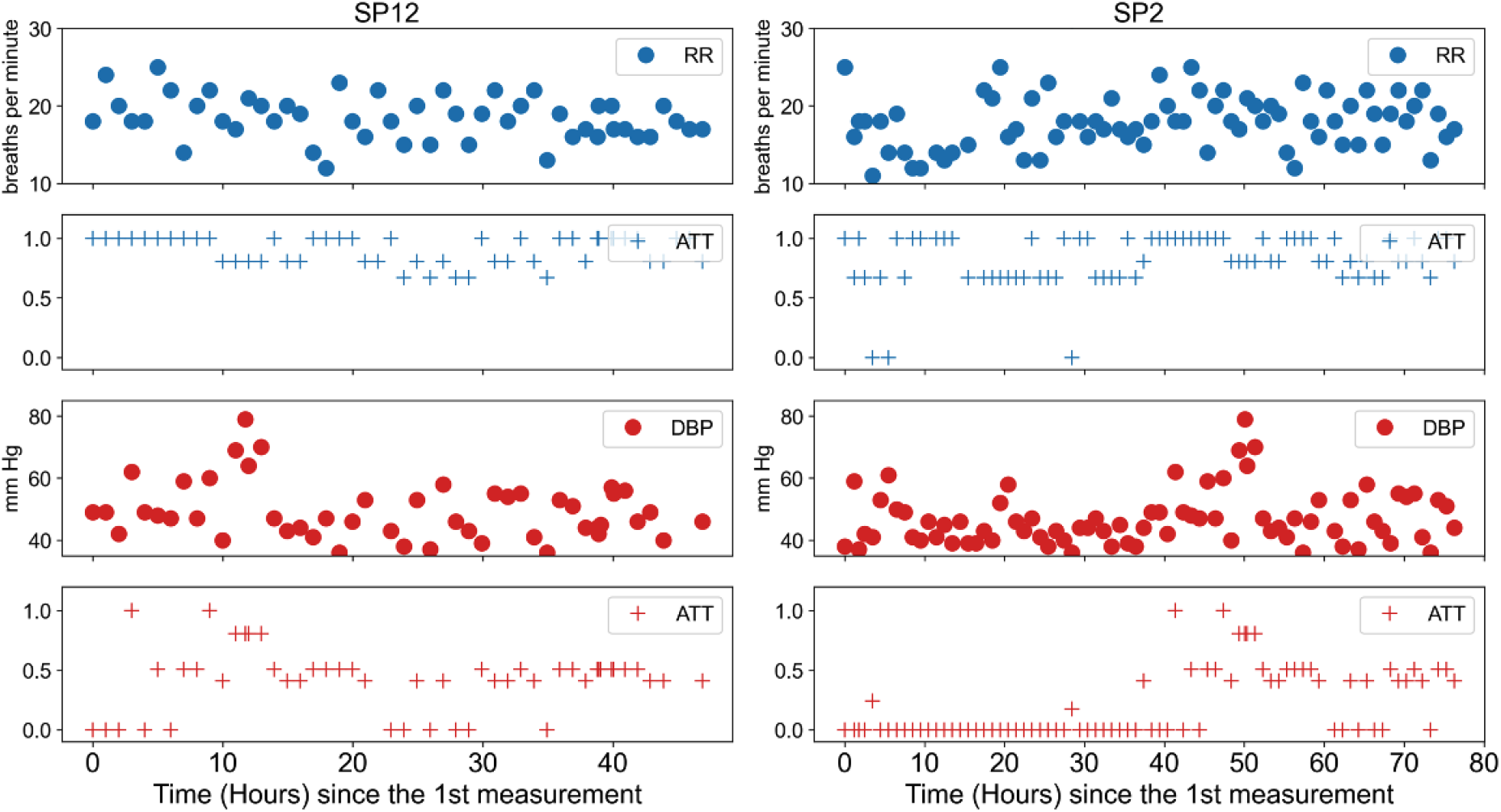
Attention scores for two samples of one patient. RR: respiratory rate; DBP: diastolic blood pressure; ATT: represents attention scores.

## V. Conclusion

In this work, we presented KIT-LSTM, a new LSTM variant that uses two time-aware gates to address irregular and asynchronous problems in multi-variable temporal EHR data and uses a knowledge-aware gate to infuse medical knowledge for better prediction and interpretations. Experiments on real-world healthcare data demonstrated that KIT-LSTM outperforms the state-of-art ML methods on continuous mortality risk prediction for critically ill AKI-D patients. In the future we will further investigate the model fairness problem and approaches to better incorporate biomedical knowledge.

## Data Availability

All data produced in the present study are available upon reasonable request to the authors

## Footnotes

1 Δ_*t*_ is repeated for every hidden dimension, thus Δ_*t*_ ∈ *ℝ*^*m*^

